# ANALYSIS OF IMMUNE ESCAPE VARIANTS FROM ANTIBODY-BASED THERAPEUTICS AGAINST COVID-19

**DOI:** 10.1101/2021.11.11.21266207

**Authors:** Daniele Focosi, Fabrizio Maggi, Massimo Franchini, Scott McConnell, Arturo Casadevall

**Affiliations:** North-Western Tuscany Blood Bank, Pisa University Hospital, Pisa, Italy; Department of Medicine and Surgery, University of Insubria, Varese, Italy; Laboratory of Microbiology, ASST Sette Laghi, Varese, Italy; Division of Transfusion Medicine, Carlo Poma Hospital, 46100 Mantua, Italy; Department of Medicine, Johns Hopkins School of Public Health and School of Medicine, Baltimore, MD, USA

**Author notes:** corresponding author: via Paradisa 2, 56124 Pisa, Italy. **Author contributions:** D.F. conceived the manuscript; F.M. analyzed the literature; S.M. provided the figure and revised the final version; A.C. and M.F. revised the final version. All authors approved the final version.

**Keywords:** SARS-CoV-2, COVID-19, Spike, convalescent plasma, viral clearance., Q493R, E484K, deletions

## Abstract

Accelerated SARS-CoV-2 evolution under selective pressure by massive deployment of neutralizing antibody-based therapeutics is a concern with potentially severe implications for public health. We review here reports of documented immune escape after treatment with monoclonal antibodies and COVID19 convalescent plasma (CCP). While the former is mainly associated with specific single amino acid mutations at residues within the receptor-binding domain (e.g., E484K/Q, Q493R, and S494P), the few cases of immune evasion after CCP were associated with recurrent deletions within the N-terminal domain of Spike protein (e.g, ΔHV69-70, ΔLGVY141-144 and ΔAL243-244). Continuous genomic monitoring of non-responders is needed to better understand immune escape frequencies and fitness of emerging variants.

## Introduction

SARS-CoV-2 Spike protein is the target of neutralizing antibody (nAb)-based therapeutics. Control of the COVID19 pandemic is being hampered by continued evolution of SARS-CoV-2, which includes mutations in the Spike protein that can affect immunogenicity and antibody-mediated neutralization. Evolutionary modeling suggests that SARS-CoV-2 strains harboring 1-2 deleterious mutations naturally exist, and their frequency increases steeply under positive selection by monoclonal antibodies (mAb) and vaccines [1]. In 2% of COVID cases, SARS-CoV-2 variants with multiple mutations occur, including in the Spike glycoprotein, which can become the dominant strains in as little as one month of persistent in-patient virus replication [2]. While mutations can occur as a natural phenomenon of SARS-CoV-2 RNA replication and editing, the pace of mutagen emergence can also be affected by small-chemical antivirals (e.g. remdesivir [3] or molnupiravir [4]). Since antibody-based therapies targeting the spike protein would also put selective pressure on SARS-CoV-2, it is reasonable to assume that widespread deployment of nAb-based therapeutics could accelerate Spike immune escape by selecting for variants resist neutralization.

Mutations that confer *in vitro* resistance to therapeutic anti-Spike mAbs have been characterized with various methods, and are informative about treatment-emergent immune escape. Deep mutational scanning (DMS) predicts protein expression, ACE2 binding, and mAb binding [5]. The method was first deployed with yeast display libraries [6], then evolved to phage display libraries (https://jbloomlab.github.io/SARS-CoV-2-RBD_MAP_clinical_Abs/) [44] and finally mammalian cell surface display [7]. nAb binding is common within the fusion peptide and in the linker region before heptad repeat (HR) region 2. The complete escape maps forecast SARS-CoV-2 mutants emerging during treatment with mAbs, and allow the design of escape-resistant nAb cocktails. Complete map of SARS-CoV-2 RBD mutations that escape bamlanivimab and its cocktail with etesevimab have been generated [8, 9].

Although DMS was also applied to polyclonal antibodies in COVID19 convalescent plasma (CCP) [10], the problem is much more complex such that it is almost impossible to identify escape mutations in CCP or vaccinee elicited sera, given the huge heterogeneity in antibody response among CCP donors and vaccinees, respectively. *In vitro*, continuous passaging of SARS-CoV-2 in the presence of a CCP unit with nAb titer >1:10^4^ led to ΔF140 Spike mutation at day 45, followed by E484K at day 73, and an insertion in the N-terminal domain (NTD): these accumulating mutations led to complete immune escape [11]. Similarly, K417N, E484K, and N501Y mutations were selected when pseudotyped SARS-CoV-2 was cultured in the presence of vaccine-elicited mAbs [12]. Although some have speculated that the large-scale use of CCP for COVID-19 could have played a role in the emergence of variants there is no evidence for such an effect and the most likely explanation for regular emergence of variants has been huge number of affected individuals since each infection case provides a natural opportunity for variant creation [13].

*In vivo*, while intra-host SARS-CoV-2 mutation development is typically very low [14], faster mutation rates (referred as “accelerated evolution”) have been found in longitudinal studies of immunodeficient patients who had persistent SARS-CoV-2 infections for several months and were treated with nAb-based therapeutics. In this study we analyze and compare the available mutational data from SARS-CoV-2 under *in vitro* and *in vivo* selection and demonstrate that mAb and polyclonal (CCP) therapies elicit different types of mutational patterns.

## Methods

We mined PubMed (which also indexes the bioRxiv and medrXiv preprint servers) for keywords related to COVID19 (“COVID19”, “SARS-CoV-2”), immune escape (“immune escape”, “treatment-emergent resistance”) and nAb-based therapeutics (“convalescent plasma”, “casirivimab”, “imdevimab”, “bamlanivimab”, etesevimab”, “sotrovimab, “regdanvimab”) both *in vitro* and *in vivo*. Clinical cases were annotated for eventual underlying immune deficiency, concurrent treatments and outcome. Figure 1 reports the study selection process according to PRISMA 2020 guidelines [15].

**Figure 1.**
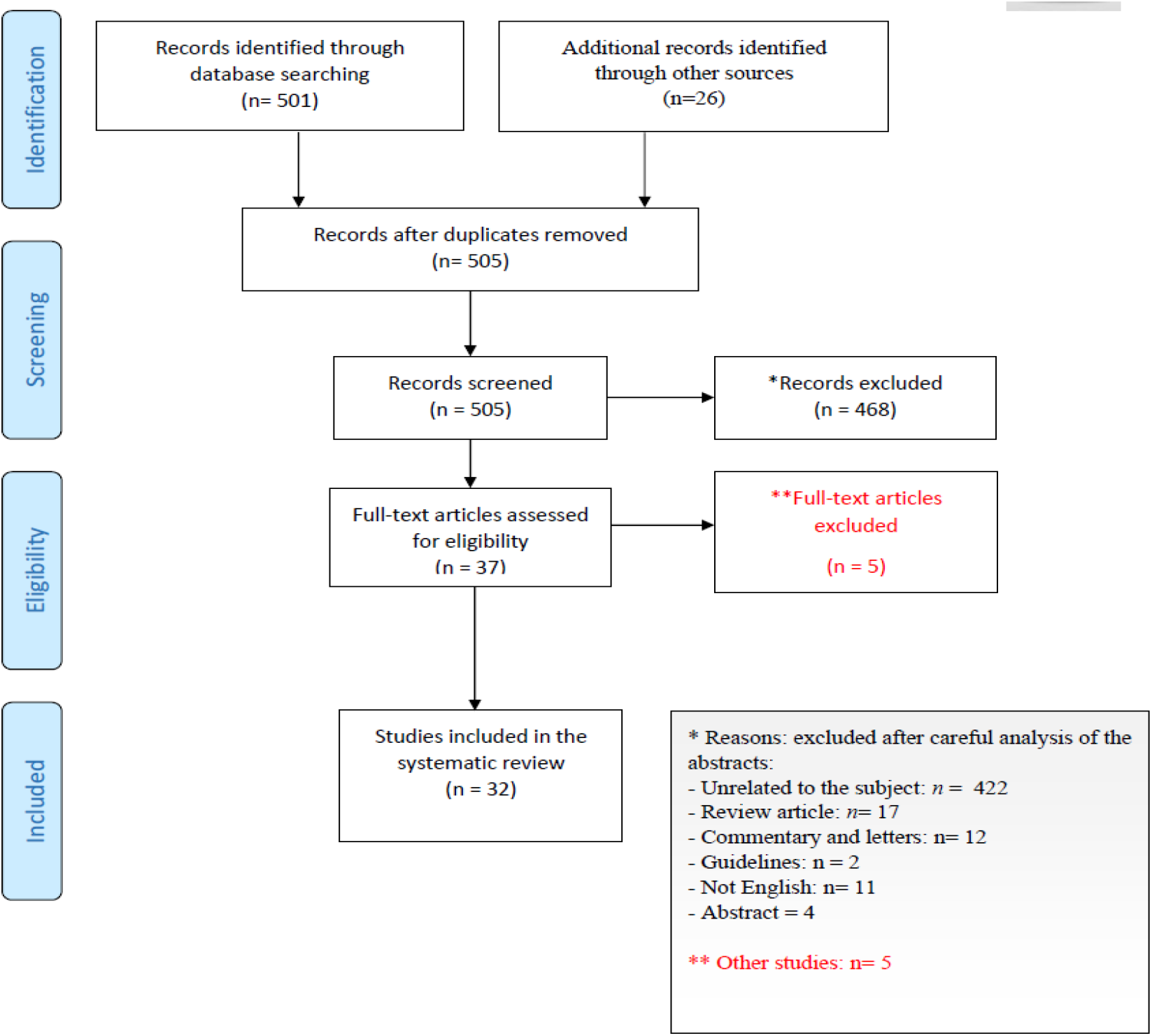
PRISMA flow diagram of study selection.

The 3D structural coordinates of the full Spike protein (PDBID 6VXX; residues 27-1252) [16] and the receptor binding domain (PBDID 7BWJ; residues 319-529) [17], solved by cryo-electron microscopy and X-ray crystallography, respectively, were used to map mutational positions of interest. Mapping on the full Spike was used to illustrate the diverse set of mutations throughout the Spike glycoprotein, while the mutations localized to the RBD were illustrated using the more complete structural model obtained through crystallography. The mutations identified in each condition of *in vivo* or *in vitro* selection were tabulated and highlighted on the structures using color coding with PyMOL [18].

## Results

Our literature search revealed 32 papers that were then manually inspected to determine whether they included relevant information that was then retrieved, evaluated and organized into Tables.

Table 1 summarizes Spike protein mutations associated with *in vitro* resistance to mAbs targeting this protein. These mutations were used to filter clinical case reports of treatment resistance for evidence of immune escape (Table 2).

**Table 1.** Spike mutations associated with clinically-approved mAb resistance *in vitro*. Mutations conferring resistance to both mAbs within the cocktail are underlined.

| manufacturer | cocktail brand name | active ingredient (brand name) | Spike mutations associated with *in vitro* resistance | ref |
| --- | --- | --- | --- | --- |
| Eli Lilly (AbCellera/Junshi) | n.a. | etesevimab (LyCoV016, CB6, JS016, LY3832479) | K417N/T, N460I, I472D, E484K, G485P, <u>Q493R/K</u> | [8, 9, 19] |
|  |  | bamlanivimab (LY-CoV555, LY3819253) | L452R, E484K, G485P, <u>Q493R/K</u>, S494P |  |
| Regeneron/Roche | REGN-COV2 (Ronapreve®) | imdevimab (REGN10987) | <u>E406W</u>, K444x, G446x | [8] |
|  |  | casirivimab (REGN10933) | <u>E406W</u>, F486x |  |
| Celltrion | - | regdanvimab (CT-P59) | n.a. | n.a. |

**Table 2.**
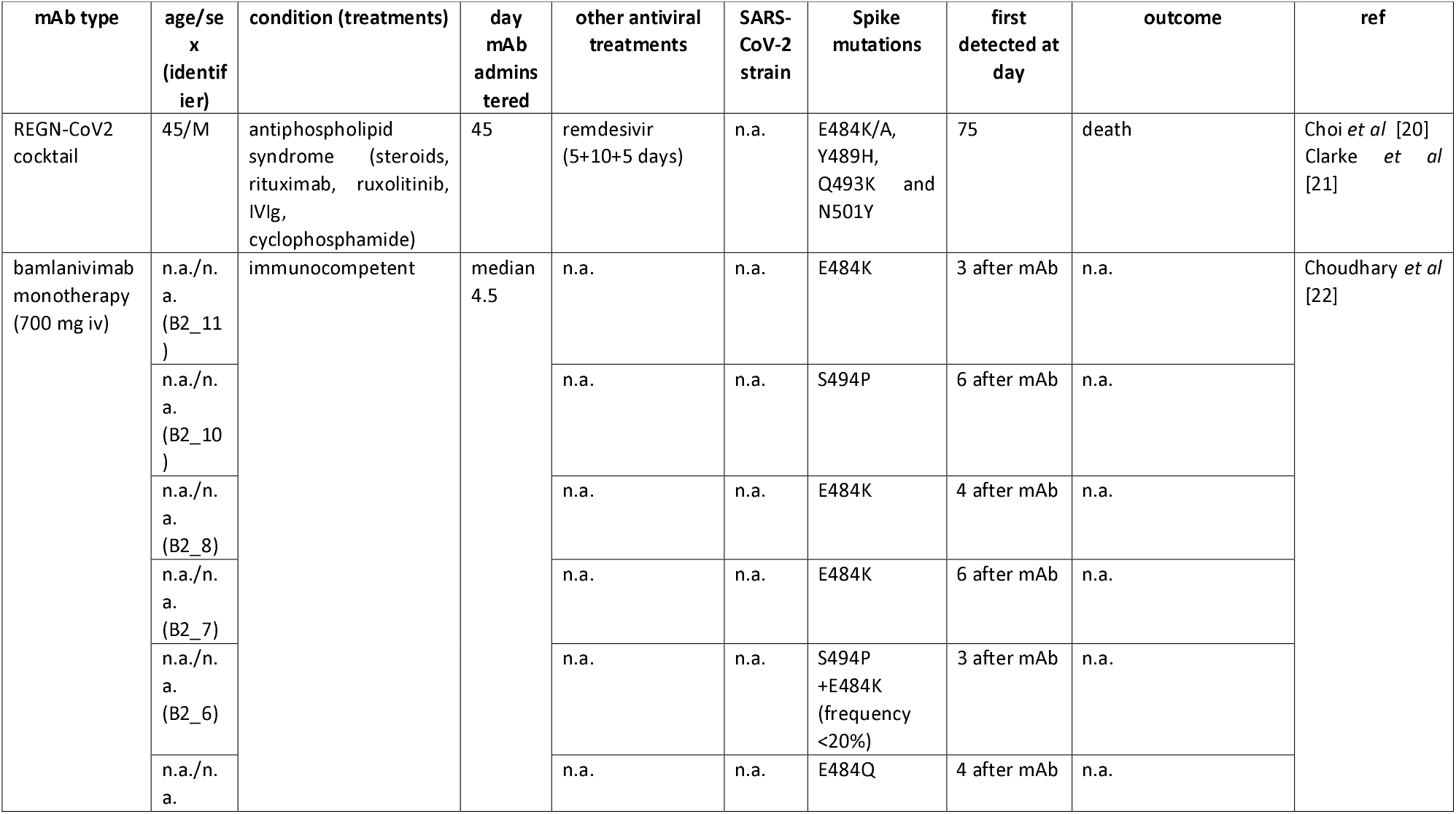
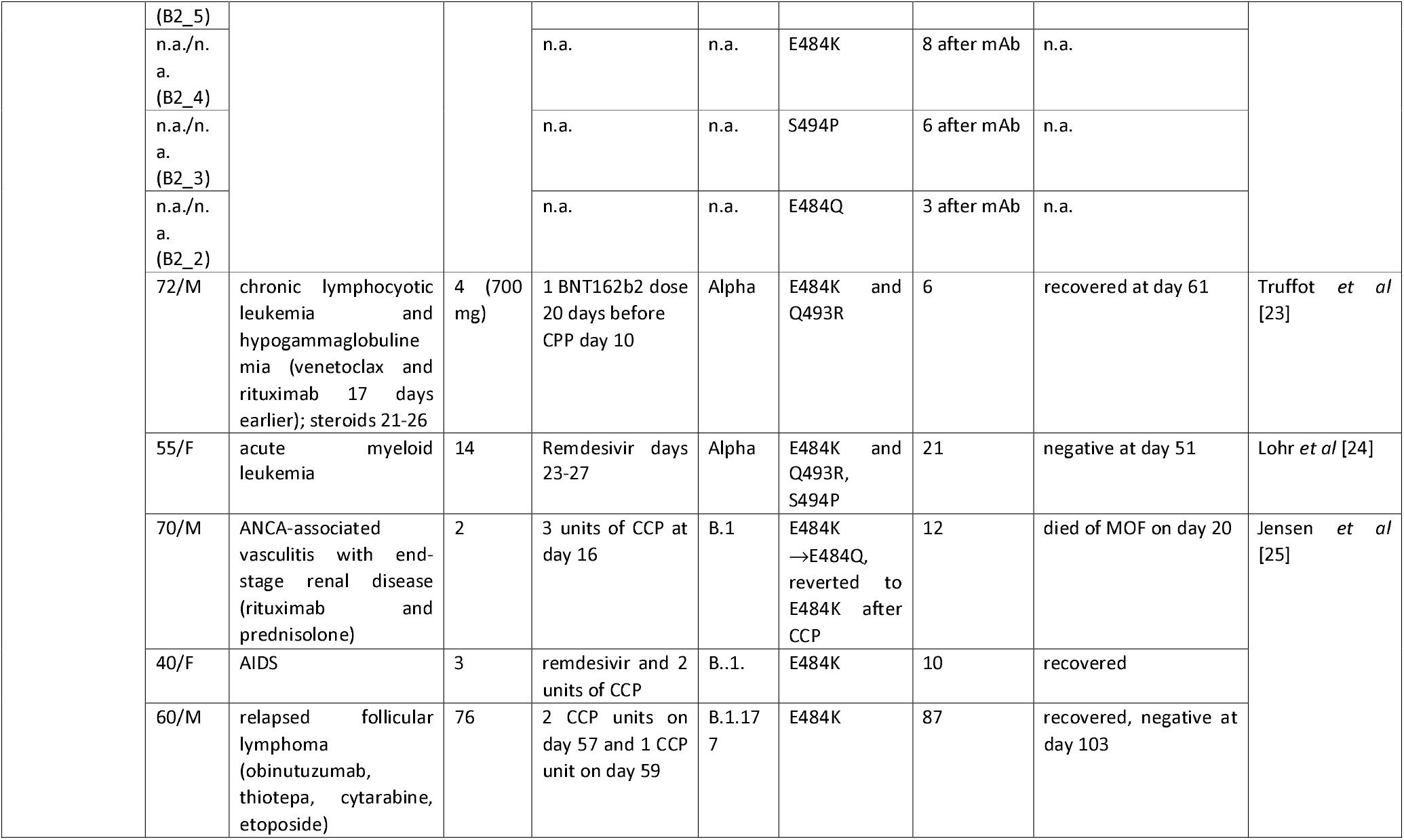
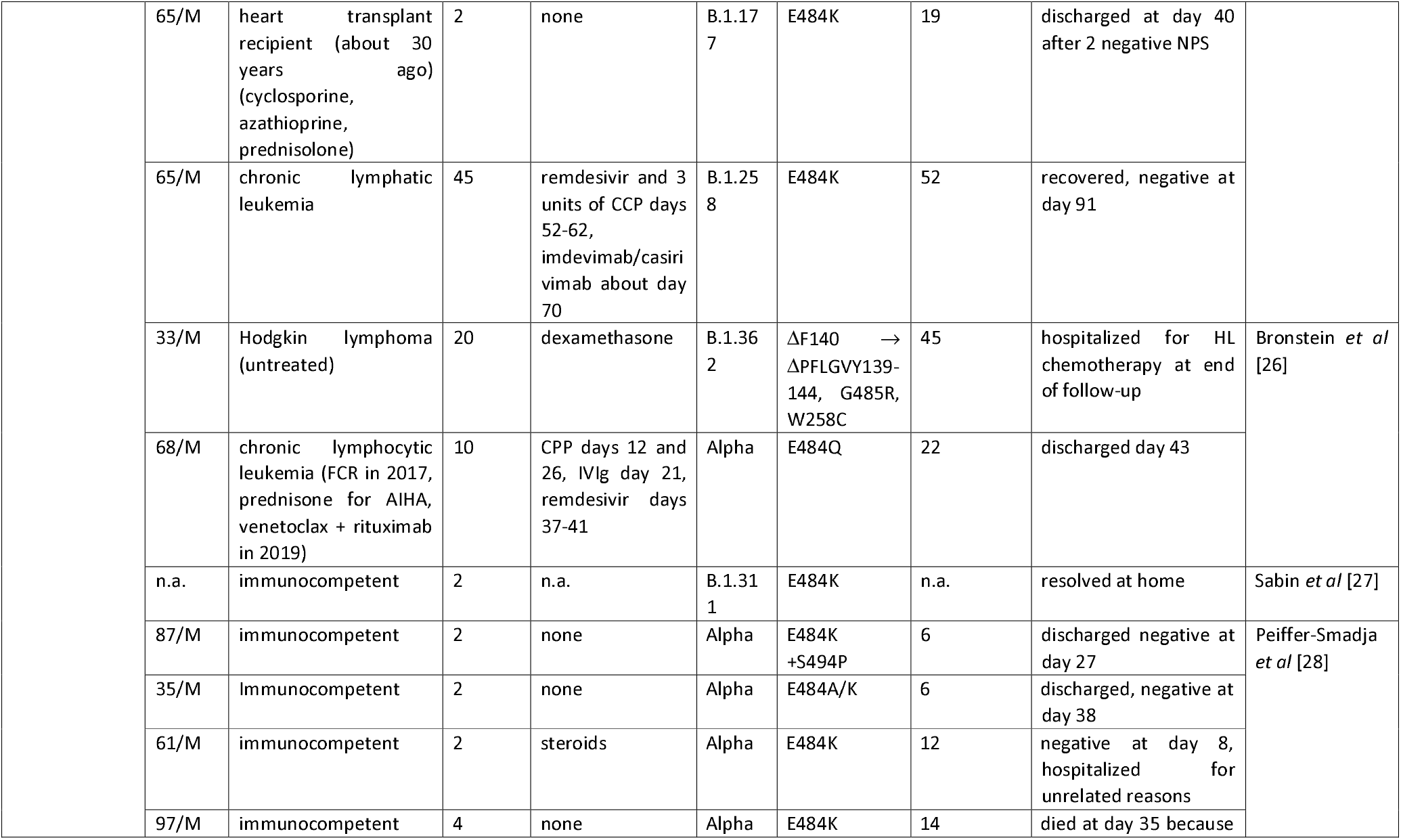
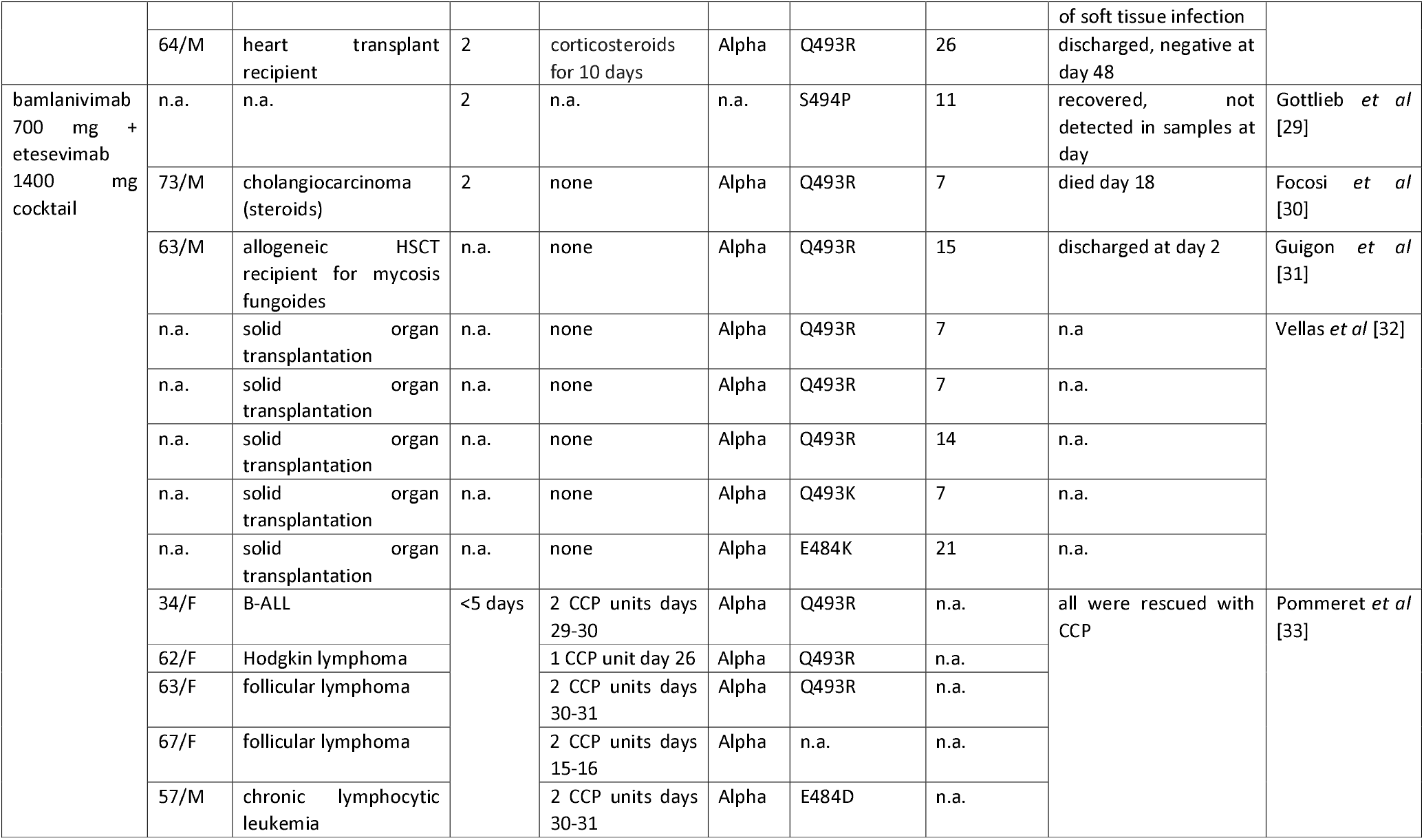
Case reports of immune escape after anti-Spike mAb treatment.

Table 3 summarizes Spike mutations found in clinical cases after CCP treatment, where, immune escape can be hypothesized to have occurred based on treatment failure, with the caveat that there is no definitive proof of immune escape due to heterogeneity of the (uncharacterized) polyclonal response.

**Table 3.** Case reports of immune escape after CCP treatment.

| age/sex<br>(identifier) | condition | CCP<br>schedule<br>(and<br>titer) | co-treatments | SARS-CoV-2<br>strain | Spike mutations | first<br>detected<br>at day | outcome | ref |
| --- | --- | --- | --- | --- | --- | --- | --- | --- |
| 71/F | chronic lymphocytic<br>leukemia and iatrogenic<br>hypogammaglobulinemia | 70 (1:60)<br>and 81<br>(1:160) | IVIg q4-6w | n.a. | ΔPFLGVYY139-145 | 49 | negative<br>NPS since<br>day 105 | Avanzato *et al* [34] |
|  |  |  |  |  | ΔLG VY141-144 | 70 (poor<br>causality) |  |  |
| 73/M | CAR-T-cell recipient | low titre<br>days 2<br>and 58 | remdesivir days 5-10,63-74<br>dexamethasone days | GH | R190K and G1124D | 13 | died day<br>74 | Hensley *et al* [35] |
|  |  |  |  |  | ΔY144, D215G, and<br>N501T | 67 |  |  |
|  |  |  |  |  | ΔH146 | 72 |  |  |
| 70/M | B-cell depletion and<br>hypogammaglobulinemia | 63, 65,<br>102 | remdesivir day 38-48, 52-62 and 91-101 | n.a. | D796H and ΔHV69-70 | 57 | died on<br>day 102 | Kemp *et al* [36] |
| 21/M | B-acute lymphoblastic<br>leukemia (CART<br>tisagenlecleucel) | 78, 103,<br>110, 123,<br>130, 137,<br>144, 158,<br>165, 172 | remdesivir (2 5-days<br>courses) | n.a. | 3 major allele variants<br>emerged between<br>days 0 and 40 with an<br>additional 4 major and<br>7 minor allele variants<br>by day 144 (ΔLGV141-143, ΔY145,<br>ΔLG VY141-144,<br>ΔNL211-212, N440K,<br>V483A, and E484Q) | 144 | positive<br>NPS at end<br>of follow-up (day<br>250) | Truong *et al* [37] |
| 50/M | kidney transplant recipient<br>(tacrolimus, steroids) | 1 | tocilizumab day 2 | B.1.369 | Q493R, ΔAL243-244<br>had ~70% frequency;<br>ΔLG VY141-144, E484K<br>and Q493K had ~30%,<br>~20% and ~10% | 21 | died on<br>day 94 | Chen *et al* [38] |
|  |  |  |  |  | frequency |  |  |  |
| 75/M | B-CLL (FCR, ibrutinib) | 2 units on day 70, 2 units on days 127-128 | remdesivir days 24-33 and 60-64 | n.a. | H49Y, ΔY144, ΔLLA241-243, ΔAL243-244, L242H, A243P, F490S, N1178N, and C1250F | 80 | still positive at end of follow-up (day 333) | Monrad *et al* [39] |
| 60/M | mantle-cell lymphoma and associated B-cell immunodeficiency (rituximab, bispecific mAb, cyclophosphamide, doxorubicin, prednisone) | 31, 122 | remdesivir day 30 and 122 | n.a. | mutations in ORF1a but not in Spike | n.a. | still positive at end of follow-up (day 156) | Baang *et al* [40] |
| 40/F | diffuse large B-cell lymphoma (CART) and hypogammaglobulinemia | high-titer day 2, 313 | IVIg, remdesivir day 2 and 313 | B.1.332 | ΔLHR244-246 and A243G | 313 (poor causation) | discharged day 324, cleared at day 335 | Nussenblatt *et al* [41] |
| 70/F (A) | follicular lymphoma (obinutuzumab-CHOP) | 23, 34, 49, 55, 56, 62, 65, 70, 73, 77, 84, 86, 90, 94, 106 | steroids | B.1.1.29 | L18F, R682Q, ΔY144 | 50 | died 5 months later | Khatamzas *et al* [42] |
| 70/M | mantle cell lymphoma (R-BAC) | 88 | darunavir/ritonavir, hydroxychloroquine, methylprednisolone, tocilizumab days 1-78, remdesivir days 45-50 and 78-87, 180-184 and 210- | B.1.1 | H69Y/P, V70G and S982A | 238 | died on day 271, still positive at day 268 | Sepulcri *et al* [43] |
|  |  |  | 214, IVIg |  |  |  |  |  |
| 40/M | autologous hematopoietic stem cell transplant due to a DLBCL | 2 doses on days ? | IVIg | B.1.128 | ΔLGV141-143 →<br>ΔLGVY141-144 | 134 | negative PCR on day 196 | Mendes-Correa *et al* [44] |

Table 4 summarizes data from reports within-host clonal evolution within immunosuppressed patients not treated with nAb-based therapeutics.

**Table 4.** Intrahost variation in Spike sequence detected in immunocompromised patients not receiving nAb-based treatments.

| age/sex<br>(identifier) | condition | antiviral<br>treatments | SARS-CoV-2<br>strain | Spike mutations | first<br>detected<br>at day | outcome | ref |
| --- | --- | --- | --- | --- | --- | --- | --- |
| 47/F | diffuse large B cell lymphoma (rituximab plus polychemotherapy) | n.a. | B.1.1.163 | Y453F, ΔHV69-70, S50L, ΔLG VY141-144, T470N, and D737G | 120 | negative PCR on day 132 | Bazykin *et al* [45] |
| 61/F | diffuse large B cell lymphoma stage IVB | remdesivir for 10 days, high-dose steroids for 7 days | B.1.1.401 | V3G, S50L, N87S, A222V, ΔLTTRTQLPPAYTN18-30 and ΔLG VY141-144 | 164 | negative PCR at day 197 | Borges *et al* [46] |
| 3/F (1) | B-cell acute lymphoblastic leukemia (chemotherapy) | n.a. | 20C | silent I410I (22792:C/A) | 27 | negative PCR at day 91 | Truong *et al* [37] |
| 2/M (3) | B-cell acute lymphoblastic leukemia | remdesivir for 5 days | 20C | V483A and E484Q<br>V70P, ΔLG V141-143, N440K | 139<br>162 | negative PCR at day 196 |  |
| 37/F | advanced HIV and antiretroviral treatment failure | dexamethasone | B.1.1.273 | E484K | 6 | negative at day 233 | Karim *et al* [47] |
|  |  |  |  | K417T and F490S | 71 |  |  |
|  |  |  |  | L455F and F456L | 106 |  |  |
|  |  |  |  | D427Y and N501Y | 190 |  |  |
| 80/M | chronic lymphocytic leukemia and hypogammaglobulinemic | remdesivir days 213-230, REGN-COV-2 day 265 | B.52 | L179 | 58 | negative PCR day 311 | Kavanagh<br>Williamson<br>*et al* [48] |
|  |  |  |  | S255F, S477N, H655Y, D1620A, ΔHV69-70 | 155 |  |  |
| 40/M | autologous hematopoietic stem cell transplant due to a DLBCL | IVIg | B.1.128 | ΔLG V141-143 → ΔLG VY141-144 |  | negative PCR on day 196 | Mendes-Correa *et al* [44] |
| n.a./n.a. | transplant recipient | remdesivir | n.a. | S13I, T95I, E484G, F490L, ΔLG VY141-144, ΔLHRS244-247, and ΔSPRRARSV680-687 | n.a. | n.a. | Weigang *et al* [49] |
| n.a./n.a. | 18 B-cell non-Hodgkin<br>lymphoma | 44% CCP<br>37% remdesivir | n.a. | n.a. | requested | n.a. | Lee *et al*<br>[50] |

Figure 2 depicts the Spike RBD mutations of concern for mAb binding detected *in vitro* and *in vivo* and the Spike mutations detected after CCP usage.

**Figure 2.**
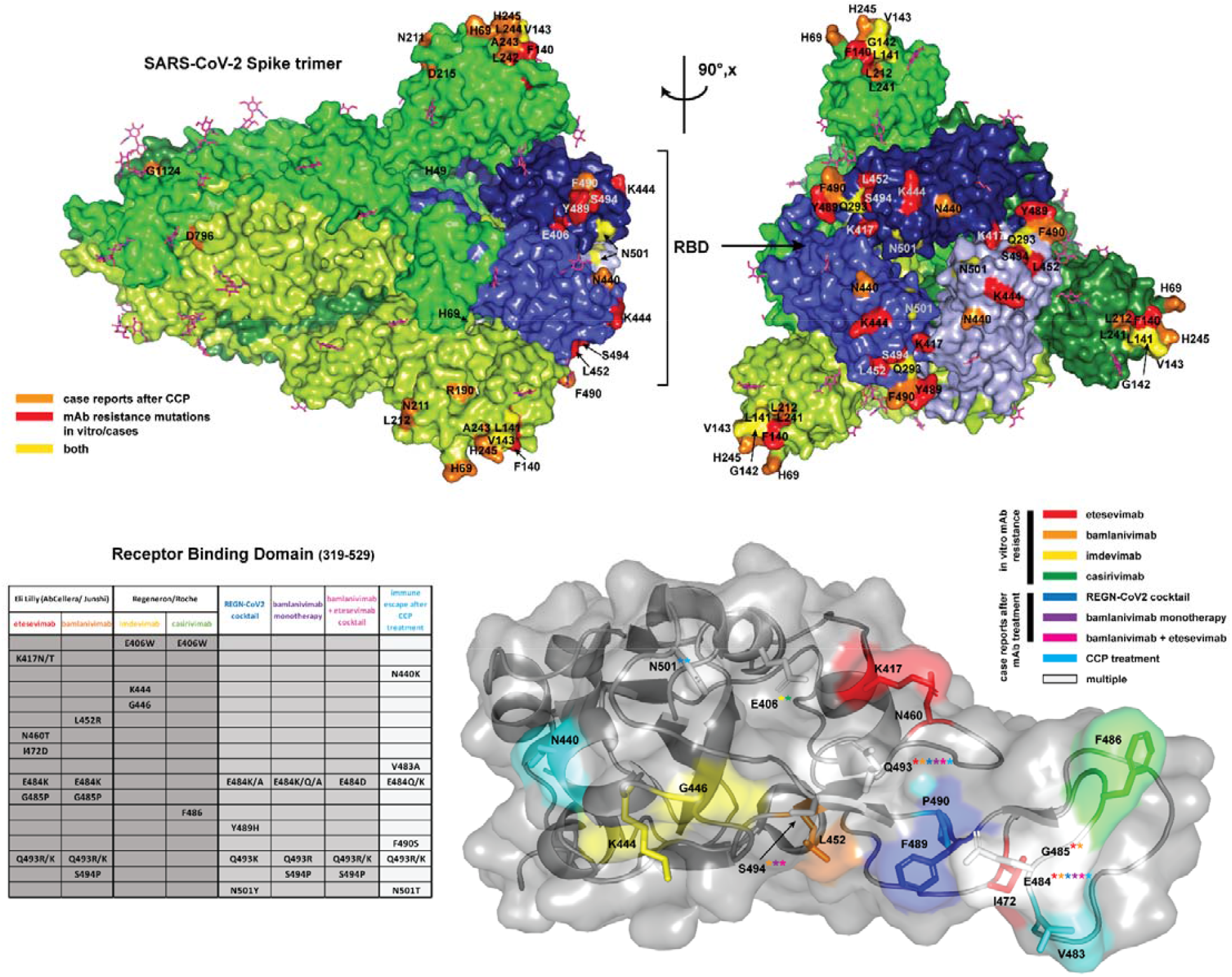
Top panel) The full SARS-CoV-2 S (spike) glycoprotein homotrimer (PDBID 6VXX) [16] in the prefusion conformation is shown in surface representation, with each spike monomer colored a different shade of green. N-linked glycosylations which were resolved in the cryo-EM map in this structure (16/22 sequons per protomer) are displayed as magenta sticks. The receptor binding domains (RBDs), in the closed state, are highlighted in 3 shades of blue corresponding to the shade of the corresponding trimer. Escape mutations from case reports of patients treated with CCP are highlighted in orange. Spike mutations associated with immune escape from clinically approved mAb treatments in vitro or from case reports are highlighted in red, while escape mutations identified in both patients who received with clinically approved mAb treatments and CCP treatment are colored yellow. The full spike is shown oriented along the long axis (left) and rotated 90 degrees to display mutations concentrated in the RBDs. Note that mutations located on unresolved loops on the cryo-EM map of the full spike are not visualized (L18, V70, Y144, Y145, D146, R246, W258, G446, N460, I472, V483, E484, G485, F486, R682, N1178 and C1250). Bottom panel) A table summarizing escape mutations localized to the RBD resulting from mAb treatments in vitro and case reports, as well as from CCP treatment. The crystal structure of single RBD domain (PBDID: 7BWJ)[17] from a more complete model (no missing loops) is displayed in surface view with the secondary structure superimposed in cartoon representation. Each escape mutation residue is highlighted by coloration according to the legend to right, and sidechains shown as sticks. In cases where a certain position corresponds to escape mutations from multiple treatments, the position is colored white and the label includes asterisks with the colors corresponding to each treatment where the escape mutation was identified. All figures were generated in PyMOL [18].

## Discussion

Escape from nAb based therapeutics provides a crucial demonstration that these immune therapies target protective antigens, which the pathogen actively evades. Hence, the emergence of neutralizing-resistant variants in individuals receiving mAb and CCP provides powerful evidence for their antiviral activity. This evidence is independent of reduction in viral load, which has been reported with mAbs given early in disease but have been an inconsistent finding in randomized controlled trials (RCT) of CCP for COVID-19 [51].

Getting frequencies for this phenomenon from case series is not possible due to the high risk of selection biases, which would yield unrealistically high frequencies. In contrast, RCTs with their control groups are the suggested reference. With bamlanivimab, there was no emergence of resistance in the patients receiving 7000 mg, but resistance was reported in patients receiving 700 mg (8 cases: 7% vs 0% with placebo) [22]. Putative treatment-emergent bamlanivimab-resistant variants were detected in 7.1% of patients (7/98) in the 700 mg group, 9.8% of patients (10/102) in the 2800 mg group, 11.3% of patients (11/97) in the 7000 mg group, 1% of patients (1/102) in the bamlanivimab and etesevimab combination group, and in 4.8% of patients (7/145) in the placebo group. The bamlanivimab monotherapy groups had a higher frequency of patients in whom a variant was detected at more than 1 time point during the viral time course (4.1% for the 700 mg group, 5.9% for the 2800 mg group, and 7.2% for the 7000 mg group) than the placebo group or the bamlanivimab and etesevimab combination group (both 0%) [29]. Apart from registration trials, the largest case series to date evaluated the impact of mAbs on the nasopharyngeal (NP) viral load and virus quasi-species of mAb-treated patients using single-molecule real-time sequencing after bamlanivimab alone (4 patients), bamlanivimab/etesevimab (23 patients) and casirivimab/Imdevimab (5 patients) [32]. To date a single case of immune escape has been reported for the non-overlapping REGN-COV2 cocktail, and accordingly hamster models and clinical trials showed no emergence of variants [52]. Since mAb therapy by definition targets only a single epitope within the RBD, it is unsurprising that escape mutations observed after *in vitro* and *in vivo* selection by these mAbs were single amino acid substitutions localized almost exclusively to the RBD (Figure 2, bottom panel; Tables 1 and 2), as expected from *in vitro* studies with single mAb, but largely prevented by non-overlapping mAb cocktails [53].

In contrast to mAb therapeutics, immune escape under CCP has not been investigated in RCTs. Hence evidences exclusively stem from case series and case reports [54], and is further complicated by exposure to multiple CCP units from different donors, each one having a polyclonal response at differing titers and affinity. Unfortunately nAb titers were very rarely determined or reported, precluding correlations between emergence of resistance and subneutralizing CCP doses. Overall, it seems that escape variants from CCP selection have not been reported as commonly nor emerged as fast. E.g., none out of 8 recipients of HSCT or CART who were treated with CCP and tested SARS-CoV-2-positive for 2 months showed significant mutations compared to the original strain [55]. Review of the Spike protein changes associated with resistance after CCP therapy reveal that most of them had in-frame amino acid deletions in a flexible region that is partially solvent exposed and forms a β strand: plasticity may contribute to the structural permissibility of the identified deletions. The NTD is a flexible region that can be affected by immune escape via either insertions (causing additional glycosylation sites [11]) or recurrently deleted regions (RDR) ΔHV69–70 (RDR1), ΔLGVY141–144 and ΔD146 (RDR2), ΔI210 (RDR3) and ΔAL243–244 (RDR4) [56] : RDR1, RDR2 and RDR4 correspond to NTD loops N2, N3 and N5, whereas RDR3 falls between N4 and N5.

Deletions of amino acids from a protein structure generally results in greater structural changes than single amino acid changes, since these reduce the size of the protein and can trigger changes that propagate through the whole structure. Furthermore, the mechanism for the emergence of deletion variants appears to be very different from the single amino acid changes that are frequent from error-prone RNA replication and could involve deletions from RNA editing. Since CCP targets a large number of epitopes in the Spike protein while mAbs target a single epitope these molecular differences parallel what is expected from their respective selection pressures in the sense that escape from polyclonal preparations requires larger antigenic structural changes than escape from mAbs. In contrast to escape mutations selected for by mAb therapy, CCP selection yields point mutations throughout the Spike protein. This reflects the vast antigenic surface area covered by the polyclonal antibodies within CCP. Escape mutations would be theoretically selected for on the basis of the most potent antibodies present in a particular CCP unit, which may vary markedly from donor to donor, which could explain the generally divergent evolution of SARS-CoV-2 in the presence of CCP. However, residues 141-144 and 243-244 are the sites of mutations or deletions in several cases, indicating these sites may offer effective escape from CCP derived from many donors, possibly by triggering a large-scale conformational rearrangement, as discussed above. As RBD binding antibodies are often neutralizing via ACE2 receptor occlusion, it is interesting that only 23% of CCP case studies identified escape mutations within the RBD (Figure 2, top panel; Table 3). This suggests that antibody binding to other sites on the Spike protein may have additional mechanisms of neutralization (i.e., by preventing conformational change after ACE2 engagement), or that additional antibody mediated immune responses (e.g., ADCC) are equally important as direct neutralization to the antiviral response to SARS-CoV-2.

Nothing can be inferred about the fitness of an emerging mutant in the absence of selective pressure, but it is of interest that one variant with the E484K mutant that emerged after bamlanivimab therapy was able to infect multiple household contacts [27]. *In vitro*, several mutants showed similar infectivity to wild type strain but resistance to different CCP donors [36]. In one instance of immune escape associated with CCP, a variant with D796H mutation manifested modestly reduced sensitivity to neutralization by CCP that was associated with reduced infectivity, which was only partly compensated by ΔHV69-70 [36]. Even if immune escape in registration trials has been a rare phenomenon, it should be considered that in the real-world practice mAbs targeting the SARS-CoV-2 Spike protein are being reserved for use in high-risk (immunocompromised) patients. Considering the huge size of a pandemic, the likelihood of immune escape becomes relevant, raising the possibility that rare variants with enhanced fitness could drive next pandemic waves. Notably, several mutations have recurred in VOC and VOIs (e.g. E484K found in Beta and Gamma, E484Q found in Delta, or ΔLHR244-246 [41] found in VOI lambda), raising the possibility that such variants emerged during treatment of patients (iatrogenic variants), but such inference will likely remain very hard to prove. E406W mutation has never been reported in GISAID, and other E406 mutations remain exceedingly rare (worldwide 318 cases of E406Q, 41 cases of E406D, and 2 cases each from USA for E406G, E406A, E406K, and 1 case of E406V out of 4,410,787 sequences deposited in GISAID as of October 25, 2021). Similarly, Q493R has only been reported in 244 sequences and Q493K in 138 sequences (source: Outbreak.info). Lack of fixation of those mutation facilitates the imputation that these require mAb selective pressure and/or effective infection control techniques in the care of those patients prevented spill over to the general population.

Within host variation (so-called “quasi-species swarm”) is a natural phenomenon which has been reported for SARS-CoV-2 in immunocompetent patients and ultimately facilitates persistence of infection. Among 33 patients having positive NPS PCR for an average of 18 days, Voloch *et al* observed a distinguishing pattern of mutations over the course of the infection mainly driven by increasing A→U and decreasing G→A signatures, including Spike mutations (V362L, T553I, H655Y, A688V, S691F, S884F, V1176F). G→A mutations are driven by RNA-editing enzyme activities typical of innate immunity [57]. Nevertheless, several covariates can facilitate immune escape.

Immunosuppression has been postulated to be an accelerator for viral evolution. Actually, Table 4 shows that very few case reports have detailed intraclonal (within-host) evolution in patients receiving immunosuppressive treatment, and, in the absence of nAb-based therapeutics, Spike mutations rarely occurred [55].

On the other hand, co-administered small chemical antivirals can be mutagenic *per se*. Remdesivir can adopt both amino and imino tautomeric conformations to base-pair with RNA bases [58]. Both amino-remdesivir:G and imino-remdesivir:C pairs could be quite mutagenic. Serial *in vitro* passages of SARS-CoV-2Engl2 in cell culture media supplemented with remdesivir selected for drug-resistant viral populations. Remdesivir triggers the selection of SARS-CoV-2 variant with a E802D mutation in the RdRp sufficient to confer decreased sensitivity to remdesivir without affecting viral fitness. The analysis of more than 200,000 sequences also revealed the occurrence of 22 mutations in Spike, including changes in amino acids E484 and N501 corresponding to mutations identified in Alpha and Beta [59]. It has been hence been proposed than nAb-based therapeutics could amplify mutations induced by remdesivir [3]. In this regard, Table 4 shows that many of the mAb- or CCP-associated mutations emerged in individuals who were or had been treated with remdesivir (but neither mAbs nor CCP), consistent with the notion that antiviral therapy could potentiate the emergence of antibody-resistance mutations.

## Conclusion

In summary, our survey of the available mutational data show that escape variants associated with mAb and CCP therapy manifest different type of mutations. For mAbs most mutations are single amino acid replacements in the RBD domain, while most variants eliciting in patients treated with CCP exhibited amino acid deletions. In fact, it is noteworthy that RBD mutations were relatively rare in CCP escape variants. Although the numbers are relatively small, which suggests cation in making generalizations, this dichotomy in geography of mAb and CCP mutations could reflect the fact that mAbs target a single epitope where the mAb-antigen interaction can be significantly altered by single amino acid changes while CCP targets many epitopes and has several mechanisms of action, such that evading polyclonal antibody immunity is likely to require much larger Spike protein structural changes. Despite the relatively small set of variants for which there is molecular data available, the large variation of molecular solutions that allow SARS-CoV-2 to escape antibody-mediated protection is striking and suggest the need for continued vigilance in genomic surveillance, especially in cases refractory to therapy.

We declare we don’t have any conflict of interest related to this manuscript.

## Data Availability

All data produced in the present work are contained in the manuscript

## Abbreviations

nAb: neutralizing antibodies
CCP: COVID19 convalescent plasma
PSM: propensity score-matched
RCT: randomized controlled trials.

